# Neurocognitive features of mild cognitive impairment and distress symptoms in older adults without major depression

**DOI:** 10.1101/2023.12.05.23299448

**Authors:** Gallayaporn Nantachai, Michael Maes, Vinh-Long Tran-Chi, Solaphat Hemrungrojn, Chavit Tunvirachaisakul

## Abstract

**Background:** Two distinct symptom dimensions were identified in older adults who did not have major depressive disorder (MDD): a) a dimension associated with mild cognitive dysfunction, and b) a dimension related to distress symptoms of old age (DSOA). It is uncertain whether previous findings regarding the features of amnestic mild cognitive impairment (aMCI) remain valid when patients with MDD are excluded.

**Objectives:** To examine, in participants without MDD, the neurocognitive characteristics of aMCI and the objective cognitive characteristics of DSOA. Neurocognition was evaluated utilizing the Cambridge Neurological Test Automated Battery (CANTAB) and memory tests.

**Results:** This research demonstrated that CANTAB tests have the capability to differentiate between aMCI and controls. The One Touch Stockings of Cambridge, probability solved on first choice (OTS_PSFC), Rapid Visual Information Processing, A prime, and the Motor Screening Task, mean latency (MOT_ML), were identified as the significant discriminatory CANTAB tests. 37.6% of the variance in the severity of aMCI was predicted by OTS_PSFC, RVP_A’, word list recognition scores, and education. Psychosocial stressors (adverse childhood experiences, negative life events), subjective feelings of cognitive impairment, and RVP, probability of false alarm, account for 40.0% of the DSOA score.

**Discussion:** When MDD is ruled out, aMCI is linked to deficits in attention, executive functions, and memory. Psychosocial stressors did not have a statistically significant impact on aMCI or its severity. Enhanced false alarm response bias coupled with heightened psychological stress (including subjective perception of cognitive decline) may contribute to an increase in DSOA among the elderly.

Keys word: depression, mild cognitive impairment, adverse childhood experiences, stress, anxiety

## Introduction

The geriatric population worldwide is expanding at a rapid rate. By 2050, it is expected to reach 22% of the total population which implies a considerable increase from the current 11% (Kanasi et al., 2016). Older age often leads to health problems including mild cognitive impairment (MCI), a condition characterized by a subjective feeling of cognitive impairment and a mild deterioration in memory functions. Nevertheless, other cognitive abilities and activities of daily living (ADL) are essentially unaffected (Petersen et al., 2001). It is estimated that around 10-15% of persons with MCI may eventually develop Alzheimer’s disease (AD), a figure approximately ten times higher than those without MCI (Anderson, 2019; Langa and Levine, 2014). aMCI has been linked to a range of behavioral and psychological symptoms, such as tension, anxiety, depressed mood, and sleep disturbances (Amrapala et al., 2023; Yatawara et al., 2018). Additional research has established an association between mood disorders and cognitive deterioration among the elderly; in particular, depression may impair neurocognitive functions (Ismail et al., 2017; McDermott and Ebmeier, 2009; Zafar et al., 2021).

The utilization of neuropsychological assessments, such as the Cambridge Neurological Test Automated Battery (CANTAB) and the Consortium to Establish a Registry for Alzheimer’s Disease, neuropsychological battery (CERAD), is critical in the evaluation of MCI and AD. Determining cognitive decline with computerized neuropsychological testing, such as the CANTAB, is a practical and precise endeavor (Ding et al., 2022). Cognitive deficits can be accurately predicted using CANTAB tests, specifically the Paired Associates Learning (PAL) task (Chandler et al., 2008) The PAL test consistently exhibits high sensitivity and specificity when differentiating clinically defined MCI from age-matched healthy controls. The application of Spatial Working Memory (SWM) to evaluate patients with MCI is further substantiated by a prior cohort study (Cacciamani et al., 2018). In a recent meta-analysis, the utility of CERAD cognitive tests in the diagnosis of MCI was demonstrated (Breton et al., 2019). The study provided evidence that the CERAD is regarded as a benchmark instrument for assessing mild cognitive impairment. In contrast to the control group, several studies have discovered that assessments, including Word List Memory (WLM), Verbal Fluency Test (VFT), and Word List Recall (WLR), exhibit adequate efficacy in identifying MCI (Tunvirachaisakul et al., 2018).

However, two significant challenges arise when attempting to evaluate MCI or aMCI. Initially, it remains uncertain whether the aforementioned findings regarding the neurocognitive characteristics of MCI in older individuals remain valid when patients with major depression are excluded. An illustration of this can be seen in the utilization of the neuropsychological tests of the CANTAB battery to evaluate depression, as demonstrated in a prior study (Egerhazi et al., 2013; Rock et al., 2014). Furthermore, Beats et al. (1996) found a significant correlation between the number of depressive episodes encountered by patients with depression and the response latency of different CANTAB tests. Consequently, it is plausible that the neurocognitive characteristics of MCI were influenced by depressive symptoms, rendering these assessments challenging to interpret in studies of MCI that did not exclude depressed patients.

Furthermore, it is possible that the existing diagnostic criteria for aMCI are excessively lenient (Maes and Tangwongchai, 2021). The question of whether aMCI is a representative sample or whether some individuals classified as aMCI may be part of the normal control sample is the subject of an ongoing debate (Maes and Tangwongchai, 2021; Tunvirachaisakul et al., 2018). Tran-Chi et al. (2023) identified two distinct dimensions among older adults who did not present with major depression in this regard. A) The first dimension, distress symptoms of old age (DSOA), comprises indications of anxiety, tension, and neuroticism; it is associated with negative life events and adverse childhood experiences (ACEs) (Tran-Chi et al., 2023). Furthermore, the subjective perception of cognitive impairment, as measured by the first item of Petersen’s criteria (Petersen, 2004), is associated with DSOA, whereas there are no associations with objective indicators of cognitive decline.

b) The second dimension is a quantitative MCI (qMCI) score that represents the severity of objective cognitive decline. This score is calculated by extracting the first principal component from the Montreal Cognitive Assessment (MoCA), the Mini Mental State Examination (MMSE), and the modified Clinical Dementia Rating (CDR) (Morris, 1993). It has been demonstrated through cluster analysis (Tran-Chi et al., 2023) that the diagnostic criteria for aMCI proposed by (Petersen, 2004) are excessively inclusive. This is because the aMCI group includes patients with DSOA who report subjective signs of cognitive impairment. Indeed, upon eliminating these DSOA subjects, a more homogeneous cohort of patients presenting with aMCI is acquired; this subset is designated mild cognitive dysfunction (mCoDy) to distinguish it from the broader MCI category.

Consequently, after excluding individuals with severe depression and DSOA, the purpose of this study is to examine the neurocognitive characteristics of aMCI, as well as the objective cognitive characteristics of DSOA. The study’s specific objectives are to identify the neurocognitive CANTAB characteristics of aMCI that are not associated with major depression or DSOA, and to determine whether DSOA is accompanied by any impairments in the CERAD or CANTAB tests.

## Materials and methods

### Participants

Participants ranging in age from 60 to 75 years were enlisted for this study. Among them were 61 aMCI cases that were referred to the King Chulalongkorn Memorial Hospital in Thailand and 59 healthy controls. Both the aMCI and control groups were enlisted from the same catchment area, which was Bangkok, Thailand. At King Chulalongkorn Memorial Hospital, subjects with aMCI were recruited at the Geriatric Clinic, the Cognitive Fitness Center unit, the Geriatric Psychiatry clinic, and the Neuroscience Center. Senior Red Cross volunteers, Health Check-up Clinics, neighborhood senior organizations, and healthy elderly caregivers of patients with aMCI who visited the Dementia Clinic comprised the control group. Patients who were diagnosed with aMCI met the Petersen criteria as outlined by Petersen (2004) and obtained a Clinical Dementia Rating (CDR) score of 0.5. These criteria consist of the detection of both subjective and objective memory impairments, the absence of dementia, and alterations in activities of daily living (ADL). Individuals with aMCI fulfilled the Petersen criteria (Petersen, 2004) and had a Clinical Dementia Rating (CDR) score of 0.5. Individuals who were diagnosed with neurological disorders such as Alzheimer’s disease, Parkinson’s disease, stroke, epilepsy, or multiple sclerosis were ineligible to take part in the study. Patients who were diagnosed with neuropsychiatric disorders were also excluded from the study, including chronic fatigue syndrome, substance use disorders, major depression, schizophrenia, bipolar disorder, generalized anxiety and post-traumatic stress disorder, obsessive-compulsive disorder, and autism spectrum disorders. Participants who had medical conditions such as cancer, chronic obstructive pulmonary disease, chronic kidney disease, metabolic syndrome, chronic inflammatory bowel disease, or HIV infection were also precluded from the study. Individuals who were unable to communicate or speak, had vision impairment or blindness (even with corrective lenses), had hearing loss, were unable to sit stably due to physical conditions such as low back pain or chronic pain, or were unable to stand or walk were not permitted to participate. Approved by the Institutional Review Board committee of the Faculty of Medicine Chulalongkorn University and the ethical committee of King Chulalongkorn Memorial Hospital (886/64), written informed consent was obtained from all subjects prior to their participation in the study.

### Study design

This study utilized a case-control design including 59 controls and 61 aMCI participants. To determine the required sample size, a power analysis was conducted, considering an effect size (f) of 0.3, alpha (α) of 0.05, power of 0.80, two groups, and four covariates. This power analysis revealed that a sample size of 90 participants would be sufficient to conduct an ANCOVA at a power of 0.8. To increase the power of our analysis and to account for potential dropouts, we included 30 more subjects, resulting in a final study sample size of 120 participants. Based on machine learning techniques performed on the same study group (Tran-Chi et al., 2023), we found two different clinical dimensions within the group of older adults and an alternative more restrictive diagnosis of aMCI, namely mCoDy (Tran-Chi et al., 2023). Using principal component analysis (PCA) performed on neurocognitive and stress-affective symptoms, we discovered two independent dimensions, namely the distress symptom of old age (DSOA) dimension, and a quantitative MCI (qMCI) dimension. The former was constructed as the first PC extracted from 6 ratings scales, namely the Perceived Stress Scale (PSS) (Wongpakaran and Wongpakaran, 2012), the State-Trait Anxiety Inventory (STAI) (Spielberger, 1983; Spielberger et al., 1971), the Thai Geriatric Depression Scale (TGDS) (Yesavage et al., 1982), the depression (HADS-D) and anxiety (HADS-A) subscale of the Hospital Anxiety and Depression Scale (Nilchaikovit, 1996), and the neuroticism trait score of Five Factor Model standardized psychometric pool of items (IPIP-NEO) (Yomaboot and Cooper, 2016). The qMCI (quantitative MCI) score was conceptualized as the first PC extracted from the total scores on the Montreal Cognitive Assessment (MoCA) (Hemrungrojn et al., 2021) and Mini-Mental State Examination (MMSE) score (Folstein et al., 1975) in a Thai version (developed by the Thai Cognitive Test Development Committee, 2002), as well as the modified Clinical Dementia Rating (CDR) score (Morris, 1993; Tran-Chi et al., 2023). In addition, cluster analysis revealed a restricted sample of subjects with cognitive dysfunctions because this ML technique removed subjects with DSOA dimension symptoms from the aMCI sample (Tran-Chi et al., 2023). As such a more restrictive aMCI subgroup was constructed (labeled mCoDy, mild cognitive dysfunction). Consequently, in the present study, we will show the results obtained on aMCI (n=61) as well as the more restricted sample mCoDy (n=52).

### Assessments

The Mini International Neuropsychiatric Interview (M.I.N.I.) (Kittirattanapaiboon, 2004) was used to make axis-1 diagnoses. All participants underwent neuropsychological testing including with the MMSE, MoCA, and CDR. In addition, we used the Cambridge Automated Neuropsychological Assessment Battery (CANTAB) (Cambridgecognition, 2022). The CANTAB tests have a proven sensitivity in detecting changes in neuropsychological performance. In the current study, we assessed key CANTAB tests (Cambridgecognition, 2022) that are relevant to aMCI namely, Delayed Match to Sample (DMS), Motor Screening Task (MOT), One Touch Stockings of Cambridge (OTS), Paired Associates Learning (PAL), Pattern Recognition Memory (PRM), Reaction Time Task (RTI), Rapid Visual Information Processing (RVP) and Spatial Working Memory (SWM). In addition, we assessed three tests of the Consortium to Establish a Registry for Alzheimer’s Disease (CERAD) (Morris et al., 1989) in a Thai translation (Tangwongchai et al., 2018), namely the Verbal Fluency Test (VFT), Word List Memory (WLM) and Word List Recall (WLR).

We conducted a structured interview to collect demographic and clinical data and which comprised questionnaires. These included the IPIP-NEO in a Thai translation (Yomaboot and Cooper, 2016), the HADS in a Thai validated translation (Nilchaikovit, 1996), the Thai versions of the State-Trait Anxiety Inventory (STAI) (Spielberger et al., 1971), and the Perceived Stress Scale (PSS) (Wongpakaran and Wongpakaran, 2012), and the validated in a Thai version of the TGDS by Poungvarin and Committee (1994). In addition, we used the Adverse Childhood Experiences (ACEs) Questionnaire (Felitti et al., 1998) in a Thai validated translation (Rungmueanporn et al., 2019) to assess five ACE dimensions, namely emotional, physical and sexual abuse, and emotional and physical neglect. We also employed the Negative Event (Hassle) Scale (Maybery et al., 2007) to assess negative life events (NLEs). The ASSIST was employed to exclude subjects with substance use, either tobacco, alcohol, cannabis, cocaine, amphetamine-type stimulants (including ecstasy), inhalants, sedatives, hallucinogens, opioids, or other drug use.

#### Statistical analysis

All statistical analyses are conducted using version 29 of IBM SPSS for Windows. We used analysis of variance (ANOVA) to investigate between-group differences in scale variables, and contingency table analyses (Χ^2^-tests) to examine associations between nominal variables. Adjusting for extraneous variables such as age, sex, and years of education, we used analysis of covariance to determine the associations between CANTAB test scores and aMCI/mCoDy compared to normal controls. To determine the best CANTAB test scores predicting aMCI/mCoDy (controls as the reference group), we utilized binary logistic regression analysis. We have accounted for potential confounding variables such as age, gender, and education in these analyses. We estimated the effect size using B (standard error, SE), Wald statistics with p-values, the Odds ratio with 95% confidence intervals (CI), the confusion matrix, and Nagelkerke pseudo-R square that was used as effect size estimate. We have employed multiple regression analysis to delineate the most significant predictors (primarily the CANTAB or CERAD test scores) on the qMCI or DSOA scores. We utilized a manual multiple regression technique in conjunction with an automated, stepwise approach, with p-values of 0.05 to enter and p=0.06 to remove. We computed standardized beta coefficients with t-statistics and p values, and essential model statistics including F values, df, and p-values. The total variance explained (R^2^) was calculated and used to estimate the magnitude of the effect. We assessed the variance inflation factor (VIF) and tolerance to check potential (multi)collinearity issues. The White and modified Breusch-Pagan tests were utilized to assess heteroskedasticity. All analyses were two-tailed, and an alpha level of 0.05 signified statistical significance.

In addition, we utilized neural networks (SPSS, Windows version 29) to determine the optimal model for predicting the outcome variables, namely the aMCI or mCoDy diagnostic classes and controls as output variables. We employed multilayer perceptron neural network models with CANTAB test scores, age, gender, and level of education as input variables. In an automated feedforward network architecture with one or two hidden layers, up to six nodes were utilized. In a batch-style training session, a maximum of 250 epochs were used to train the models. The termination criterion was established based on the failure of successive steps to further reduce the error term. The error, relative error, and percentage of misclassifications were calculated, and the predictive precision of the models was estimated using the confusion matrix of the holdout sample. The importance chart depicts the significance and relative prominence of the input variables.

Using SmartPLS (Ringle et al., 2015), the causal relationships between CANTAB and CERAD variables, age, sex, and education (used as input variables), and DSOA versus qMCI symptom dimensions were evaluated. When the outer and inner models satisfied predetermined quality criteria, a comprehensive PLS analysis was performed. To determine the validity of the model, several criteria were examined. First, all loadings on the latent vectors must be greater than 0.66 at a p <0.001 significance level. In addition, the Standardized Root Mean Square Residual (SRMR) should be less than 0.08 to indicate an adequate model fit. Furthermore, all latent vectors should demonstrate satisfactory composite reliability rho A values (> 0.7) with an average variance extracted (AVE) exceeding 0.50. Confirmatory Tetrad Analysis must verify that the factors have been specified adequately as reflective models. Using the Heterotrait-Monotrait ratio (HTMT) to evaluate discriminant validity should be sufficient (HTMT scores <0.9). The model’s reproducibility was evaluated using the Q^2^ predict values (PLSPredict).

## Results

### Socio-demographic and clinical data

**Table 1** shows the socio-demographic and clinical data of the aMCI group (n=61), and healthy controls (n=59). There were no significant differences in gender, BMI, and marital status between the groups. There was a minor difference in age between both groups and subjects in the aMCI group had a lower number of education years as compared with controls. In any case, we allowed for the effects of age, sex, and education in all multivariate analyses. The aMCI group exhibited significant lower scores on various neurocognitive tests, including the MoCA, MMSE, VFT, WLM, and WLR as compared with controls. There were no significant differences in IPIP-NEO neuroticism, HADS-A, HADS-D, STAI, ACE neglect, total NLE, and NLE money+health scores among both study groups. The PSS and TGDS scores were higher in aMCI patients as compared with controls.

**Table. 1.**
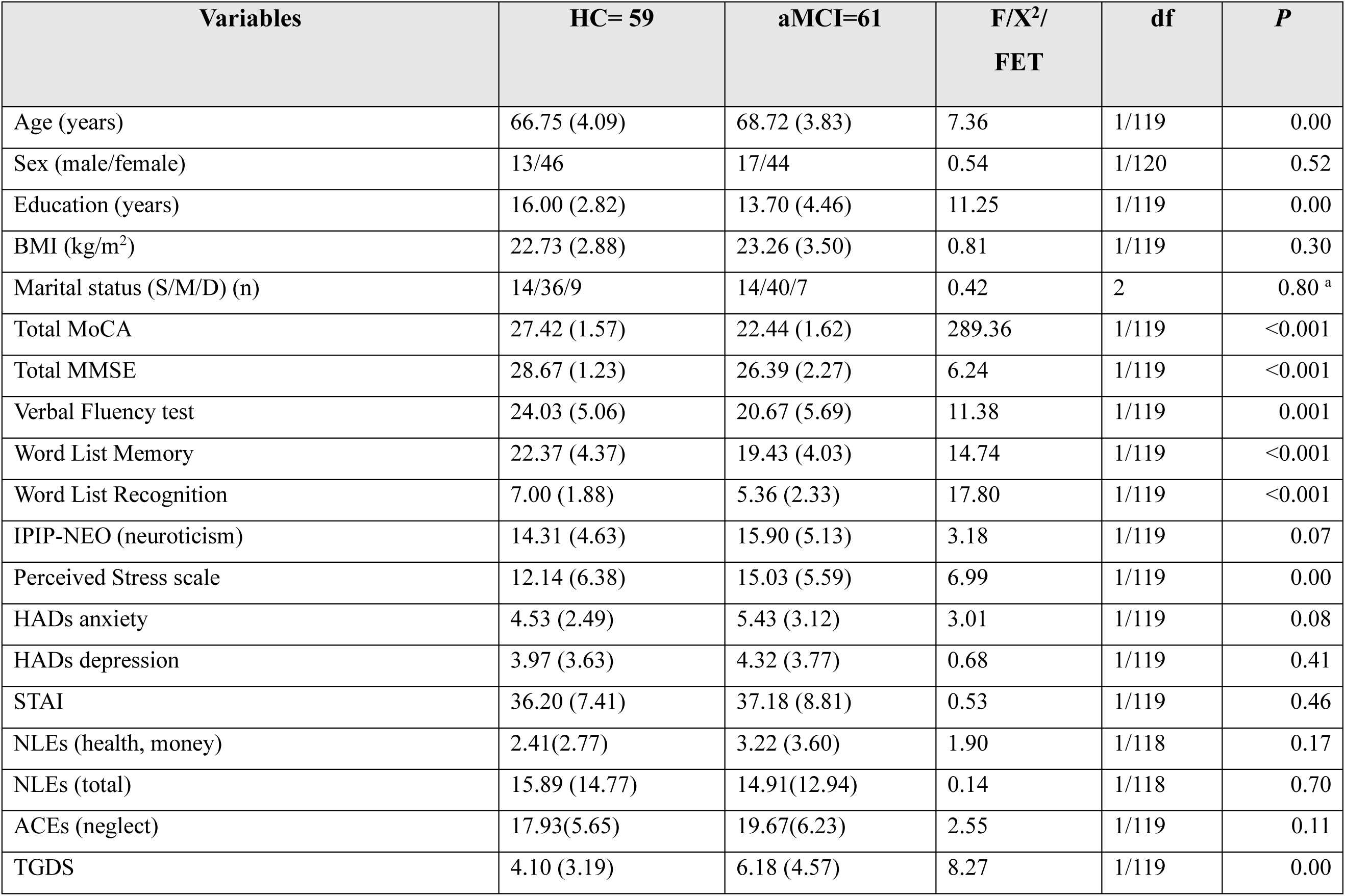
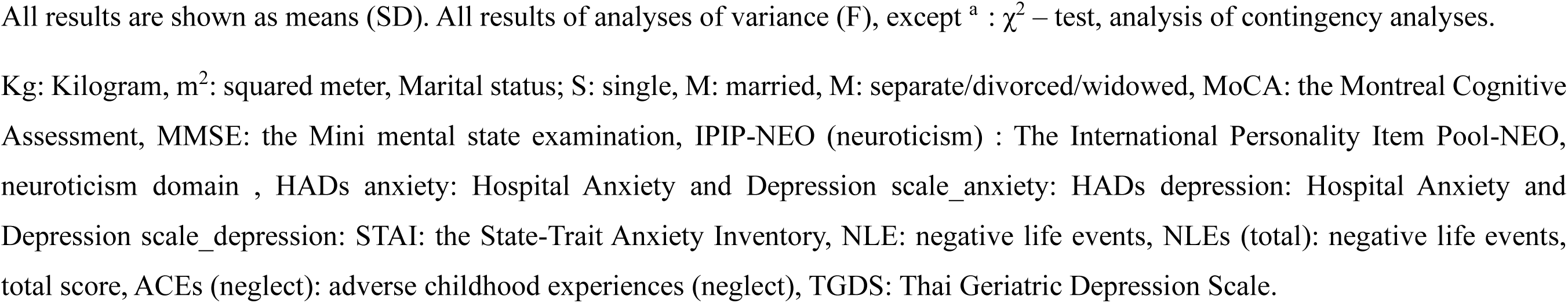
Characteristic clinical data of subjects with amnestic mild cognitive impairment (aMCI) and healthy controls (HC)

### CANTAB test scores predicting aMCI

We have employed binary logistic regression analysis using the key CANTAB tests as explanatory variables to predict aMCI (the dependent variable), while allowing for the effects of the variables age, sex, and education years. **ESF. Table 2** Shows that there are only five key CANTAB tests that were significantly associated with aMCI, namely OTS_PSFC (Nagelkerke R^2^ = 0.256), PRM_PCD (Nagelkerke R^2^ = 0.206). RVP_Á (Nagelkerke R^2^ = 0.230), MOT_ML (Nagelkerke R^2^ = 0.207). and MOT_SDL (Nagelkerke R^2^ = 0.219). The clustered bar graphs in Figure 1a and 1b show the mean test scores of the 28 key CANTAB tests in aMCI and HC.

**Table 2.** Results of binary logistic regression analysis with amnestic mild cognitive impairment (aMCI) as dependent variable and healthy controls (no aMCI) as reference group.

| Regression Number | Explanatory Variables | B | SE | Wald | P | OR | 95% CI |
| --- | --- | --- | --- | --- | --- | --- | --- |
| aMCI | Male sex | 1.254 | 0.543 | 5.34 | 0.021 | 3.501 | 1.210-10.147 |
|  | OTS_PSFC | -0.852 | 0.259 | 10.83 | <0.001 | 0.426 | 0.257-0.708 |
|  | RVP_A' | -0.713 | 0.229 | 9.66 | 0.002 | 0.490 | 0.312-0.768 |
| mCoDy | OTS_PSFC | -0.694 | 0.266 | 6.83 | 0.009 | 0.500 | 0.297-0.841 |
|  | RVP_A' | -0.576 | 0.260 | 4.89 | 0.027 | 0.562 | 0.337-0.937 |
|  | MOT_ML | 0.514 | 0.248 | 4.29 | 0.038 | 1.672 | 1.028-2.719 |
|  | Education | -0.153 | 0.067 | 5.23 | 0.022 | 0.858 | 0.753-0.978 |

mCoDy: mild cognitive dysfunction (a subgroup of aMCI subjects), OTS\_PSFC: One Touch Stockings of Cambridge, probability solved on first choice, MOT\_ML: Motor Screening Task, mean latency, RVP\_A': Rapid Visual Information Processing, A prime.

We have employed multivariable binary regression analysis to delineate the most important predictors of aMCI using the CANTAB tests scores, age, sex, and education. **Table 2**, regression #1 shows that OTS_PSFC, RVP_ Á and male sex were the best predictors of aMCI (Χ^2^=28.83, df=3, p<0.001, Nagelkerke=0.289, accuracy=66.1%). Allowing for the effects of the three CERAD tests, showed that OTS_PSFC, RVP_ Á, RVP_MDL and WLR best predicted aMCI versus healthy controls (Χ^2^=41.24, df=4, p<0.001, Nagelkerke=0.408, accuracy=72.2%). We performed a third binary logistic regression using the restricted aMCI subgroup (namely the mCoDy group) as dependent variable. Table 2, regression #3 shows the outcome of this analysis; OTS_PSFC, RVP_ Á, MOT_ML and education best predicted mCoDy versus healthy controls (Χ^2^=32.29, df=4, p<0.001, Nagelkerke=0.351, accuracy=71.1%).

**Table 3** displays the outcomes of a neural network analysis (NN#1) that investigated the effects of input variables on the prediction of output variables (aMCI and healthy controls). Age, sex, education, and selected CANTAB test scores (those that were significant in ESF, Table 2, and those with p values < 0.12) were entered as input variables. In this NN#1 model, the concealed layer activation function was hyperbolic tangent, while the output layer activation function was softmax. The training consisted of two hidden layers, with three units in layer 1 and two in layer 2. Training substantially decreased the cross-entropy error, indicating enhanced trend generalization. The consistency of the percentage of incorrect predictions across training, testing, and holdout samples suggests that model overfitting is not present. In the holdout sample, the cross-validated precision of the model was sensitivity = 56.5% and specificity = 81.0%. Figure 3 depicts the importance chart with all input variables’ normalized importances. RVP_ Á, MOT_ML, PRM_PCD, and OTS_PSFC were identified as the most significant input variables by the NN#1 model, followed by DSM_PC4, education, RVP_PFA, and age.

**Table 3.** Results of neural networks (NN) with amnestic mild cognitive impairment (aMCI) and healthy controls (HC) as output variables and the Cambridge Neurological Test Automated Battery (CANTAB) variables as input data.

|  | **Models** | **NN#1** | **NN#2** |
| --- | --- | --- | --- |
| **Input layer** | Number of units | 13 | 13 |
| **Hidden layers** | Number of hidden layers | 2 | 2 |
|  | Activation function | Hyperbolic tangent | Hyperbolic tangent |
|  | Number of units in hidden layer 1 | 3 | 3 |
|  | Number of units in hidden layer 2 | 2 | 2 |
| **Output layer** | Output variables | aMCI and HC | mCoDy and HC |
|  | Number of units | 2 | 2 |
|  | Activation function | Softmax | Identity |
| **Training** | Error term | 32.705 (cross entropy) | 11.392 (sum of squares) |
|  | % incorrect predictions | 32.1% | 28.6% |
| **ROC curve** | Area under the ROC curve | 0.801 | 0.781 |
| **Testing** | Error term | 13.390 (cross entropy) | 4.242 (sum of squares) |
|  | % incorrect predictions | 25.0% | 30.4% |
| **Holdout** | % incorrect predictions | 33.3% | 17.9% |
|  | Classification | Sensitivity=56.5%,<br>specificity=81.3% | Sensitivity=80.0%<br>specificity=83.3% |

mCoDy: mild cognitive dysfunction (a more pure subgroup of aMCI subjects),

Table 3, model NN#2 depicts a second neural network with the restricted group mCoDy and healthy controls as output variables and the selected CANTAB test scores, age, sex and education as input variables. As described above and in Long et al. (2023), this class derived from cluster analysis is a more restricted aMCI class because individuals with DSOA were excluded. In this model, the activation functions for the concealed layer and output layer were hyperbolic tangent and identity, respectively. The model was trained using a configuration with two hidden layers, with three units in the first hidden layer and two units in the second. The holdout sample’s cross-validated precision was 82.1% accurate, with a sensitivity of 80.0% and a specificity of 83.3%. **Figure 1** depicts the relevance chart displaying the normalized importances of the input variables. The model ascribed the greatest predictive power to OTS_PSFC, RVP_A’, DSM_PCS, and PRM_PCD, with education, MOT_ML, and PAL_FMAS trailing behind. RVP_PFA, sex, and age, on the other hand, demonstrated a very low predictive power for mCoDy.

**Figure 1.**
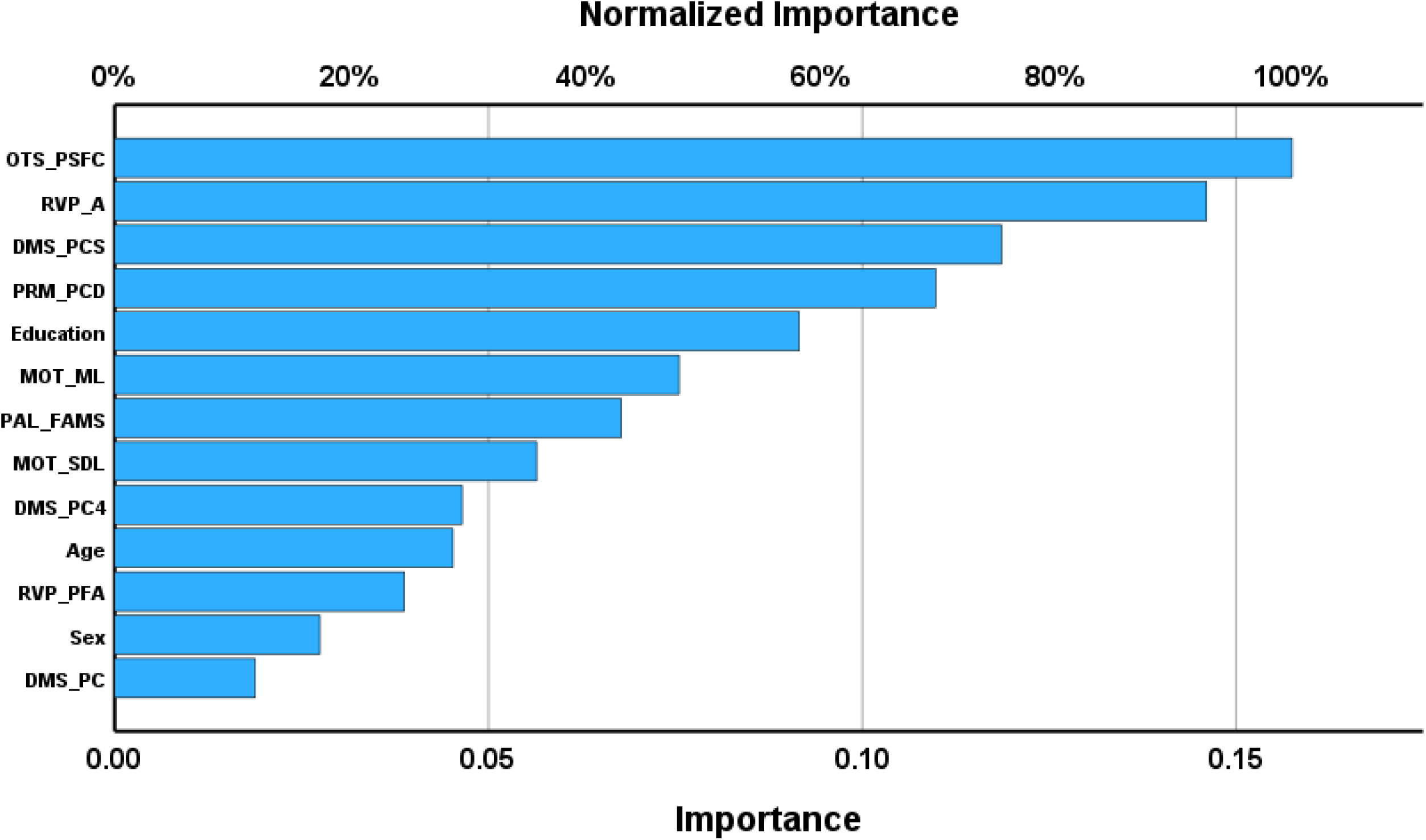
Results of neural network analysis with amnestic mild cognitive impairment and normal controls as output variables and selected key Cambridge Neuropsychological Test Automated Battery (CANTAB) test scores as input variables. OTS_PSFC: One Touch Stockings of Cambridge, probability solved on first choice, RVP Á: Rapid Visual Information Processing, A prime, DMS_PCS: Delayed Match to Sample, percent correct, PRM_PCD: Pattern Recognition Memory, percent correct delayed, MOT_ML: Motor Screening Task, mean latency, PAL_FAMS: Paired Associates Learning, first attempt memory score, MOT_SDL: Motor Screening Task, standard deviation of the mean latency, DMS_PC4: Delayed Match to Sample, percent correct 4 second delay, RVP_PFA: Rapid Visual Information Processing, probability of false alarm, DMS_PC: Delayed Match to Sample, percent correct.

### Prediction of the qMCI and DSAO latent vector scores by cognitive test results

**Table 4** shows the results of multiple regression analyses with the qMCI and DSOA scores as dependent variables and the key CANTAB tests (and VFT, WLM, and WLR in the case of mCoDy score; and Petersen item 1 in the case of the DSOA score) and age, sex, education, ACE and NLE variables as additional explanatory variables. Model #1 shows that 37.6% of the variance in the qMCI score was explained by OTS_PSFC, WLM, education and RVP_ Á (all inversely associated). **Figure 2** shows the partial regression of the qMCI score on the OTS_PSFC score. Model #2 shows an alternative model, after inclusion of the VFT test score, whereby 37.1% of the variance in qMCI is explained by OTS_PSFC, education, WLM, and VFT (all inversely associated). In both regression analyses, no significant effects of NLE, ACE, sex, or age could be established. Model #3 shows the outcome of the multivariate regression of the DSOA score on the significant predictors, namely NLE health+money, ACE neglect, education, RVP_PFA and Petersen item 1 (all positively associated). **Figure 3** shows the partial regression of the DSOA score on RVP_PFA score.

**Figure 2.**
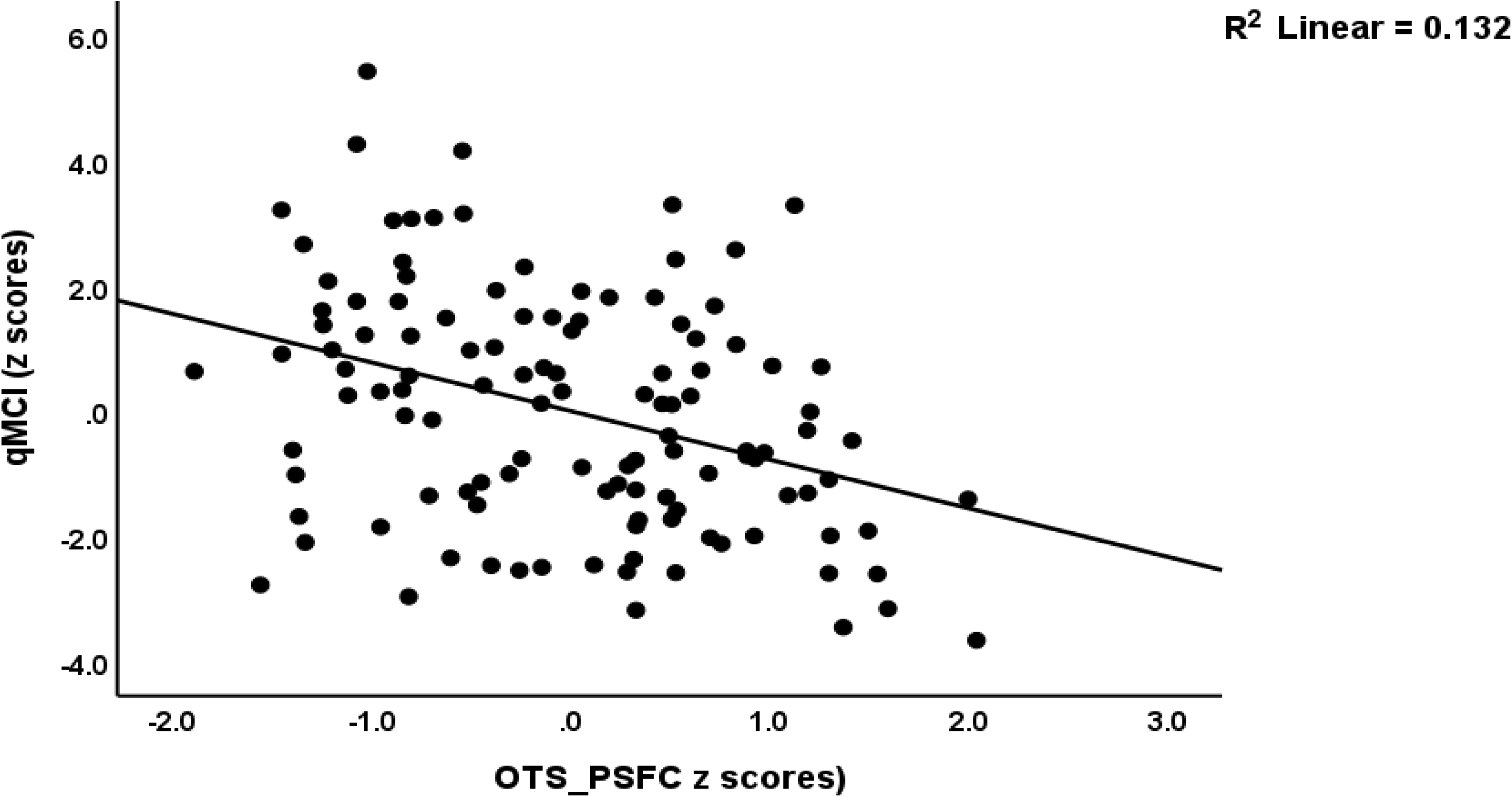
Partial regression of the quantitative mild cognitive impairment score (qMCI) on the One Touch Stockings of Cambridge (OTS), probability solved on first choice (OTS_PSFC) (adjusted for age, sex and education)

**Figure 3.**
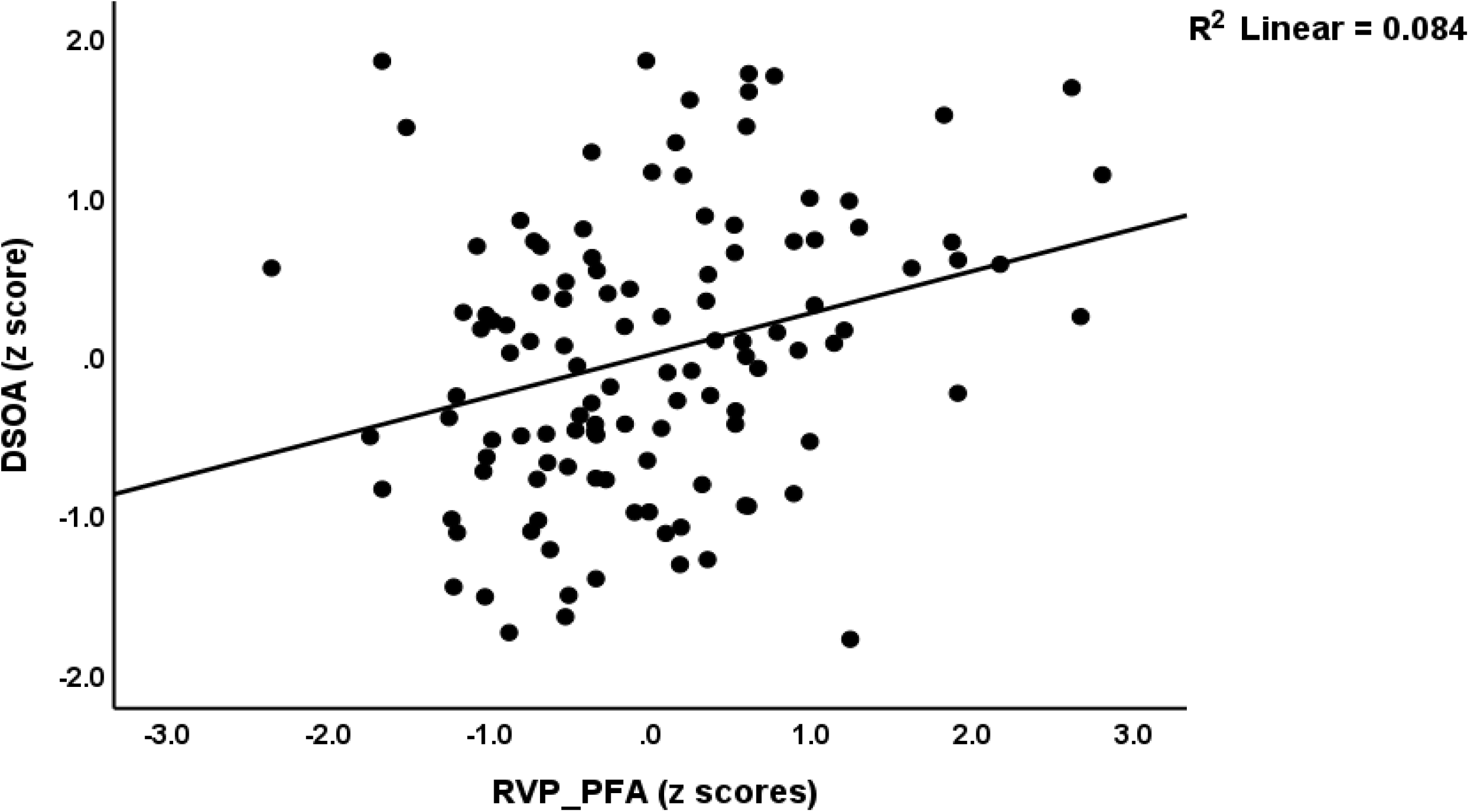
Partial regression of the distress symptoms of old age (DSOA) score on the Rapid Visual processing (RVP) probability of false alarm (PFA) score (adjusted for age, sex and education)

**Table 4.**
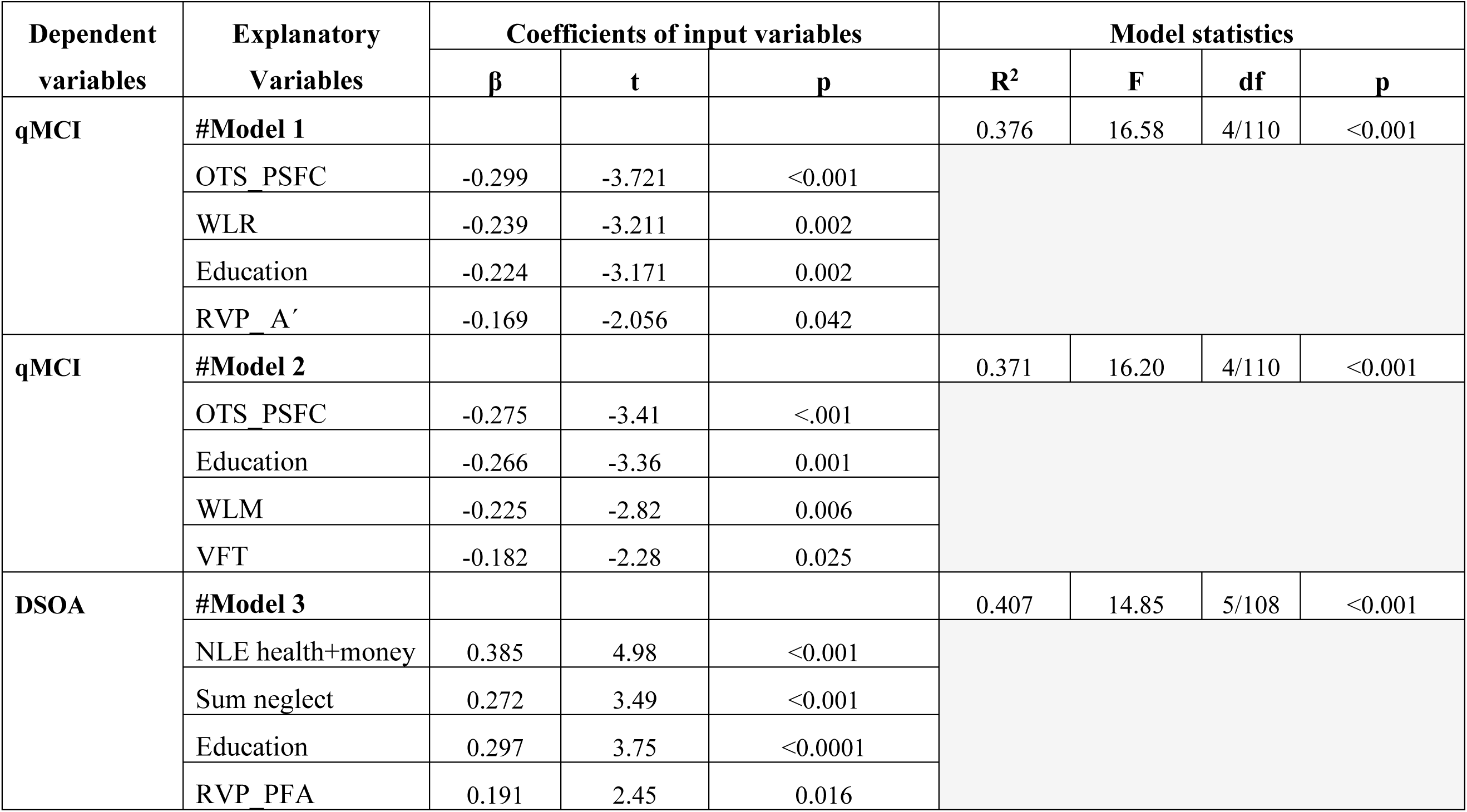
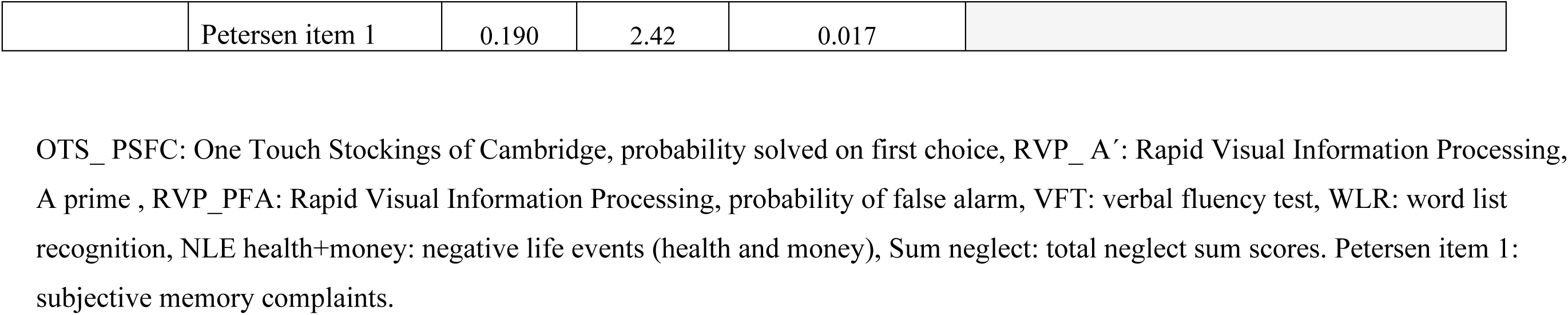
Results of multiple regression analyses with the quantitative scores of mild cognitive dysfunction (qMCI) or distress symptoms of old age (DSOA) as dependent variables, and the key Cambridge Neurological Test Automated Battery (CANTAB) Test scores as dependent variables.

| Dependent variables | Explanatory Variables | Coefficients of input variables |  |  | Model statistics |  |  |  |
| --- | --- | --- | --- | --- | --- | --- | --- | --- |
| | | $\beta$ | t | p | R<sup>2</sup> | F | df | p |
| qMCI | #Model 1 |  |  |  | 0.376 | 16.58 | 4/110 | <0.001 |
|  | OTS PSFC | -0.299 | -3.721 | <0.001 |  |  |  |  |
|  | WLR | -0.239 | -3.211 | 0.002 |  |  |  |  |
|  | Education | -0.224 | -3.171 | 0.002 |  |  |  |  |
|  | RVP A' | -0.169 | -2.056 | 0.042 |  |  |  |  |
| qMCI | #Model 2 |  |  |  | 0.371 | 16.20 | 4/110 | <0.001 |
|  | OTS PSFC | -0.275 | -3.41 | <.001 |  |  |  |  |
|  | Education | -0.266 | -3.36 | 0.001 |  |  |  |  |
|  | WLM | -0.225 | -2.82 | 0.006 |  |  |  |  |
|  | VFT | -0.182 | -2.28 | 0.025 |  |  |  |  |
| DSOA | #Model 3 |  |  |  | 0.407 | 14.85 | 5/108 | <0.001 |
|  | NLE health+money | 0.385 | 4.98 | <0.001 |  |  |  |  |
|  | Sum neglect | 0.272 | 3.49 | <0.001 |  |  |  |  |
|  | Education | 0.297 | 3.75 | <0.0001 |  |  |  |  |
|  | RVP PFA | 0.191 | 2.45 | 0.016 |  |  |  |  |

|  |  |  |  |  |
| --- | --- | --- | --- | --- |
|  | Petersen item 1 | 0.190 | 2.42 | 0.017 |

OTS\_ PSFC: One Touch Stockings of Cambridge, probability solved on first choice, RVP\_ A': Rapid Visual Information Processing, A prime , RVP\_ PFA: Rapid Visual Information Processing, probability of false alarm, VFT: verbal fluency test, WLR: word list recognition, NLE health+money: negative life events (health and money), Sum neglect: total neglect sum scores. Petersen item 1: subjective memory complaints.

### Results of PLS-path analysis

Figure 4 displays the PLS model after removing the non-significant input variables (e.g., sex and age) and non-significant paths (e.g., from DSOA to qMCI). The final outcome variables entered in the model were the DSOA and qMCI latent vectors. The former was extracted from HADS-A, HADS-D, PSS, STAI, GDS, and neuroticism scores, and the latter was extracted from the total MoCA and MMSE scores and CDR scores. Direct predictors were three CANTAB test scores (selection based on the abovementioned results of neural networks and multiple regression analysis and feature reduction), namely OTS_PSFC, RVP_ Á and RVP_PFA, Petersen item 1 (entered as a dummy variable), education (years), NLE health+money, and one latent vector extracted from VFT, WLM and WLR test scores (labelled “memory”). We also entered one formative model, namely a composite built using emotional abuse and neglect and physical neglect (labelled “ACE”). Complete PLS analysis performed using 5.000 bootstrap samples showed that the construct reliability and convergence validities of the three reflective latent vectors were adequate with AVE values > 0.599 and rho_A > 0.7, while the SRMR of 0.054 indicated an adequate model fit. The outer loadings were all > 0.66 at p<0.0001. CTA showed that the three latent vectors were not mis-specified as reflective models. All Q^2^ values exceeded zero, indicating replicability of the model. We found that 36.8% of the variance in the qMCI score was explained by the CERAD “memory” latent vector, education, OTS_PSFC and RVP_ Á. On the other hand, 42.4% of the variance in the DSOA latent variable score was explained by ACE, NLE health+money, education, Petersen item 1, and the RVP_PFA score.

**Figure 4.**
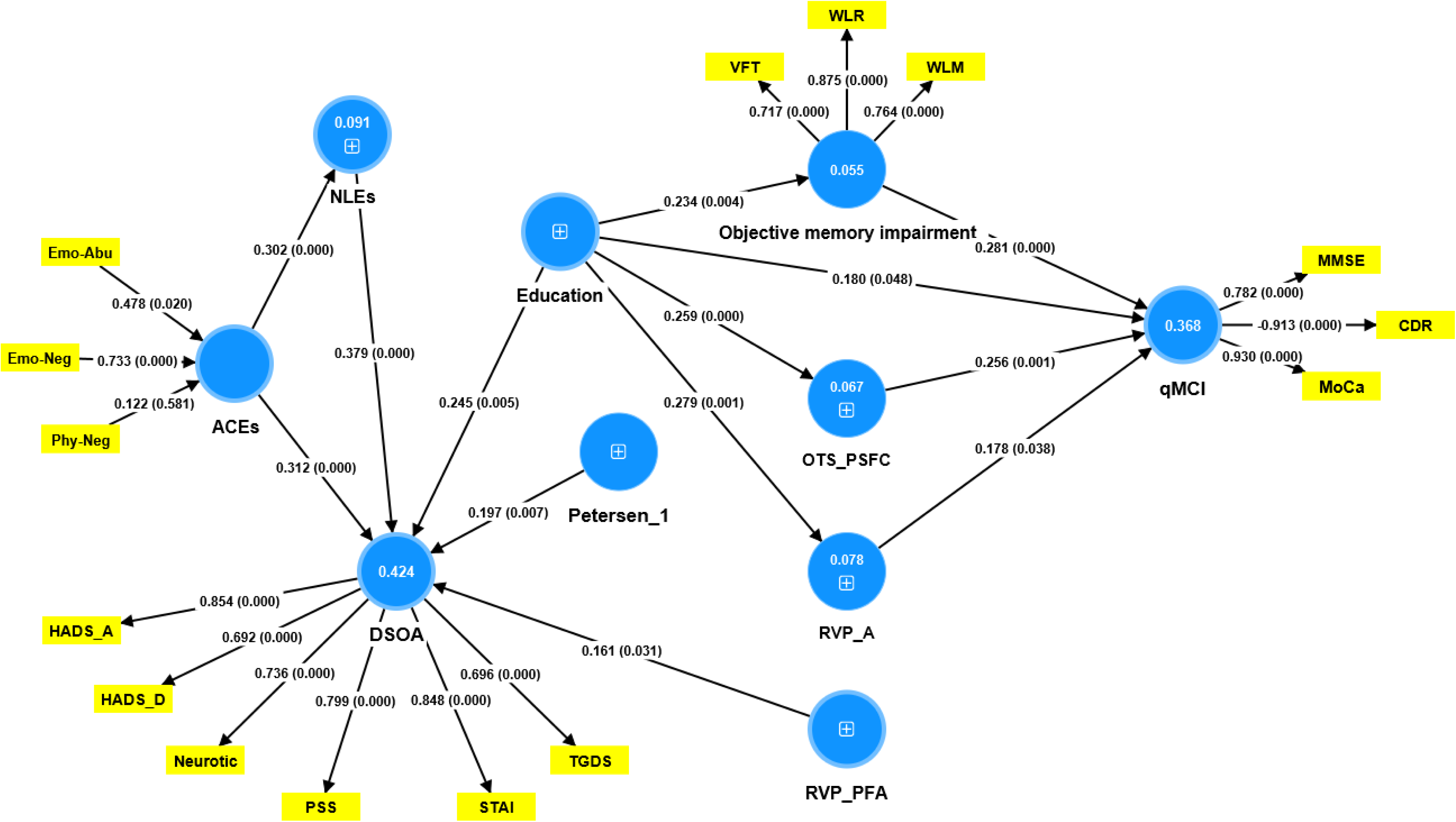
Results of partial least squares analysis. We entered two outcome variables, namely the quantitative mild cognitive impairment (qMCI) factor, and the distress symptoms of old age (DSOA) factor. Objective memory disorders was conceptualized as a factor extracted from three memory tests. Adverse childhood experiences (ACEs) was conceptualized as a factor extracted from three ACE subtypes. All other predictors were entered as single indicators. The explained variance is shown as figures in blue circles. The outer model shows the loadings on the factors and p-values. Paths are shown as path coefficients with exact p values. Emo-Abu: emotional abuse, Emo-Neg: emotional neglect, Phy-Neg: physical neglect, HADs-A: Hospital Anxiety and Depression scale-Anxiety, HADs-D: Hospital Anxiety and Depression scale-Depression, Neurotic: The International Personality Item Pool-NEO, neuroticism domain, PSS: Perceived Stress scale, STAI: the State-Trait Anxiety Inventory, TGDS: Thai Geriatric Depression Scale, VFT: Verbal Fluency test, WLR: Word List Recognition, WLM: Word List Memory, MMSE: the Thai-Mini mental state examination, CDR: the modified Clinical Dementia Rating, MoCA: the Montreal Cognitive Assessment.

In this PLS model, there was no significant association between the DSOA and qMCI scores (p=0.577). There were no significant effects of “memory” (p=0.382), OTS_PSFC (p=0.318) and RVP_ Á (p=0.901) or any of the other CANTAB tests on the DSOA factor score. There were no significant effects of NLE health+money (p=0.169), ACE (p=0.591) and RVP_PFA (p=0.485) on the qMCI latent variable score. Increasing education differently affected both dimensions, namely protecting against increasing qMCI, but contributing to increased DSOA scores.

## Discussion

### CANTAB tests and aMCI or mCoDy

The first significant finding of this study is that RVP_ Á, RVP_MDL, OTS_PSFC, and MOT_ML were among the neuropsychological CANTAB tests that distinguished aMCI or the more restrictive group mCoDy from controls. Furthermore, neural network analysis demonstrated that PRM_PCD possessed some additional ability to differentiate aMCI.

Previous research showed that RVP_Á scores were lower in multi-domain aMCI and AD patients compared to controls (Broadhouse et al., 2021). Several CANTAB tests, including RVP, differed significantly between MCI and control subjects, according to a prior systematic review (Sabahi et al., 2022). The RVP test evaluates attention and psychomotor speed as well as the capacity to maintain visual attention and detect stimuli. The RVP_Á key test assesses the subject’s ability to identify target sequences by calculating the median response latency on trials in which the subject provided accurate responses. Conversely, RVP_MDL assesses the median response time, and both critical evaluations distinguish attention (Cambridgecognition, 2022).

In a prior investigation, Levy-Gigi et al. (2011) identified a notable disparity in the outcomes of OTS tests. Individuals with MCI exhibited a distinct pattern in the mean number of attempts at faultless solutions and the mean proportion of correct solutions compared to the control group. The OTS_PSFC test computes the aggregate count of evaluated trials in which the subject select the accurate response on their initial attempt, encompassing all evaluated trials (Cambridgecognition, 2022). The OTS test score may serve as an assessment tool for executive functions, encompassing problem-solving, working memory, mental flexibility, planning, strategic thinking, response inhibition in aMCI or AD (Cambridgecognition, 2022; Chamberlain et al., 2011; Levy-Gigi et al., 2011; Swainson et al., 2001). It is noteworthy to mention that the frontal lobe of the brain is where executive functions are predominantly executed (Alvarez and Emory, 2006).

Prior research (Broadhouse et al., 2021) found that individuals with AD and multi-domain aMCI had significantly lower MOT_ML test scores than the control group. The MOT test results enable one to evaluate the reaction time, movement time, vigilance and sensorimotor deficit (Cambridgecognition, 2022). The MOT_ML test calculates the mean latency for a subject to correctly respond to the stimulus on the screen during the assessed trials.

PRM can predict the cognitive status of MCI patients, according to multiple investigations (Campos-Magdaleno et al., 2021; Nathan et al., 2017). The PRM test evaluates visual pattern recognition memory in the context of a 2-choice forced discrimination paradigm; its design is intended to be sensitive to the medial lobe.

Numerous experimental studies have demonstrated the noteworthy efficacy of video-game training in ameliorating cognitive functions among individuals diagnosed with mild cognitive impairment (MCI). According to a study by Leelavanichkul and Hemrungrojn (2013), video games improved executive function and cognitive speed, as demonstrated by enhanced results on the OTS and PAL CANTAB tests. Furthermore, when applied to individuals with MCI, game-based neurofeedback demonstrated favorable outcomes, including enhanced attention and working memory, as evaluated by the SWM test (Jirayucharoensak et al., 2019). However, the current study failed to identify any statistically significant variations in the measurement of SMW between subjects with aMCI and the control group, even though this test is frequently linked to MCI (Sabahi et al., 2022).

### Prediction of the severity of MCI

The second main finding of this study was that the combined effects of OTS_PSFC, RVP_Á, WLM, WLR, VFT, and/or education predicted a quantitative score of mild cognitive dysfunction by a substantial margin (around 37% of the variance). As reviewed in the Introduction, earlier studies on cognitive functions in MCI focused primarily on the PAL test results (Barnett et al., 2016; Junkkila et al., 2012; Zhuang et al., 2021). However, the current research revealed the significance of executive functions (the OTS_PSFC test results) combined with episodic memory, semantic memory, verbal fluency, recognition memory, and working memory (PRM, RVP, VFT, WLM, and WLR test results) as predictors. Previously, Tran-Chi et al. (2023) demonstrated that RVP_MdL, SWM_Ber, and DMS_Cor account for 29.0% of the variance in a factor extracted from the MoCA and MMSE test scores and the diagnosis MCI (which is quite like the qMCI score that we utilized in the present study). This finding suggests that the RVP test is, in fact, the most replicable determinant of the severity of MCI.

Memory assessments, including those conducted by the CERAD, have been shown in a prior meta-analysis to be rather specific for MCI (Breton et al., 2019). As previously demonstrated by Tunvirachaisakul et al. (2018), the VFT, WLM, and WLR may in fact be effective at detecting MCI. By utilizing the CERAD-subtest scores, including those from the VFT, Modified Boston naming test, WLM, true recall, WLR, and constructional praxis, a noteworthy distinction was obtained between individuals with MCI and those who were asymptomatic, yielding an area under the ROC curve of 0.862 (95% CI = 0.816–0.908) (Aguirre-Acevedo et al., 2016). Prior research indicates that the CERAD total scores and various individual CERAD measures have the potential to effectively identify MCI (Dos Santos et al., 2011; Kraaij et al., 2002; Paajanen et al., 2010; Sala et al., 2017).

It is noteworthy that within our study sample, which excluded participants with major depression and BPSD, neither ACE nor NLEs had a significant impact on aMCI, mCoDy, or the quantitative qMCI score. However, ACEs and NLEs significantly predicted DSOA. An increased risk of MCI has been linked to heightened psychological stressors, according to several studies (Avery et al., 2023; Franks et al., 2021). In addition, negative social interactions may increase the risk of MCI and cognitive decline in older adults (Tran-Chi et al., 2023; Wilson et al., 2015). Furthermore, individuals with MCI may exhibit higher physical neglect scores compared to the control group (Wang et al., 2016). However, after excluding patients with major depression, our study revealed that there are no such associations between the severity of aMCIs and ACEs or NLEs. Therefore, it is highly likely that the correlation between stressors and MCI can be attributed to the presence of depression; consequently, the association between MCI and psychological stressors found in previous studies may represent a spurious correlation.

### The DSOA score is predicted by false alarm

The third significant discovery of this study is that the DSOA score, which encompasses distress, emotional, and neuroticism scores, is influenced not only by ACEs and NLEs, but also by a subjective perception of cognitive impairments and the RVP_PFA test scores. In this respect, it is interesting to note that a subjective cognitive decline, which is a feature of DSOA, has been linked to ACEs, such as sexual, physical, and psychological abuse (Brown et al., 2022). The RVP_PFA test score is a particular cognitive predictor for DSOA that is not linked to MCI or the severity of MCI. RVP_PFA detects and reacts to quick variations in the environment; it is calculated as the proportion of incorrect alarms in consecutive presentations compared to the total of incorrect alerts plus accurate rejections (Cambridgecognition, 2022). False alarms are crucial in the signal detection theory (Feldman, 2021). Stressors have the tendency to alter response bias in highly anxious individuals (Hoskin et al., 2014). Moreover, anxiety is linked to decreased ability to distinguish between hazardous and neutral stimuli when exposed to unclear threat cues (Ozturk et al., 2023) As such, the DSOA score’s severity can be attributed to a combination of psychosocial stressors and false alarms in detecting distress and anxiety symptoms.

## Limitations

There were certain limitations in the study. Conducting the study in a metropolitan area such as Bangkok may produce findings that are not applicable to elderly individuals residing in the rural regions of Thailand. Hence, it is imperative to replicate the findings of the current study in rural regions and across other cultural or national contexts.

## Conclusions

This study demonstrated that some CANTAB tests had the ability to differentiate between individuals with aMCI or the more specific category of mCoDy, and control subjects. The crucial CANTAB tests include RVP_Á, RVP_MDL, OTS_PSFC, MOT_ML, and PRM_PCD. The severity of MCI was primarily determined by a combination of two CANTAB tests (OTS_PSFC and RVP_Á) along with CERAD memory tests. The findings indicate that MCI, after removing major depression and DSOA, is linked to deficits in attention, executive functioning, and memory. Crucially, when we removed older adults with major depression, BPSD and DSOA from the analysis, we found no significant impact of psychosocial stressors on MCI or its severity. On the other hand, psychosocial stresses such as ACEs and NLEs, and the subjective perception of cognitive deficits are predictors of DSOA. Additionally, RVP_PFA suggests that false alarms can play a role in the development of DSOA, while there is no evidence of objective cognitive deterioration in the latter domain.

Subsequent investigations should reassess the connections between MCI or its severity and neurocognitive test outcomes, biomarkers, and AD onset. This should be done by eliminating patients with major depression and by accounting for DSOA. By doing this, researchers will be able to define the specific features of (a)MCI more clearly.

## Supporting information

ESF table 1-2, ESF figures1-3

## Data Availability

All data produced in the present study are available upon reasonable request to the authors

## Acknowledgments

Not applicable.

## Ethical approval and consent to participate

The research project (IRB no.886/64) received approval from the Institutional Review Board (IRB) at Chulalongkorn University’s ethics board in Bangkok, Thailand. This approval is in accordance with the International Guidelines for the Protection of Human Research Participants, as mandated by the Declaration of Helsinki, The Belmont Report, CIOMS Guidelines, and the International Conference on Harmonization in Good Clinical Practice (ICH-GCP).

## Declaration of interest

The authors declare no conflicts of interest.

## Funding

The study was supported by the 90^th^ Anniversary of Chulalongkorn University Scholarship under the Ratchadaphisek Somphot Fund Batch#52/2 (GCUGR1125652003D), and the Ratchadaphisek Somphot Fund (Faculty of Medicine), MDCU (Batch #GA65/34), The scholarship from graduate affair, Faculty of Medicine, Chulalongkorn University, Thailand, to GN. This research is funded by Thailand Science research and Innovation Fund Chulalongkorn University (HEA663000016) to MM.

## Credit authorship contribution statement

Gallayaporn Nantachai: data curation, investigation, project administration, first draft writing; Michael Maes: design, methodology, conceptualization, statistical analysis, visualization, writing, review, supervision; Vinh-Long Tran-Chi: data curation, investigation, project administration; Solaphat Hemrungrojn: editing; Chavit Tunvirachaisakul: design, methodology, review, and editing. All authors approved the submitted draft of this papers.

## Availability of data

The corresponding author MM will provide the data file used in the present study upon receiving an appropriate request once the authors have fully utilized the data.

## Notes

### Competing Interest Statement

The authors have declared no competing interest.

### Funding Statement

This study was funded by by the 90th Anniversary of Chulalongkorn University Scholarship under the Ratchadaphisek Somphot Fund Batch#52/2 (GCUGR1125652003D), and the Ratchadaphisek Somphot Fund (Faculty of Medicine), MDCU (Batch #GA65/34), The scholarship from graduate affair, Faculty of Medicine, Chulalongkorn University, Thailand, to GN. This research is funded by Thailand Science research and Innovation Fund Chulalongkorn University (HEA663000016) to MM.

### Author Declarations

Approved by the Institutional Review Board committee of the Faculty of Medicine Chulalongkorn University and the ethical committee of King Chulalongkorn Memorial Hospital (886/64), written informed consent was obtained from all subjects prior to their participation in the study.

