## Supplementary material for "Neurocognitive features of mild cognitive impairment and distress symptoms in older adults without major depression": ESF table 1-2, ESF figures1-3

**ELECTRONIC SUPPLEMENTARY FILE (ESF)**

**Running title:** Mild cognitive Impairment and distress symptoms in old age.

Gallayaporn Nantachai^*^, Michael Maes^*^, Vinh-Long Tran-Chi, Solaphat Hemrungrojn, Chavit Tunvirachaisakul.

ESF, Table 1. Results of the principal symptoms of distress symptoms of old age components (PC_DSOA) analysis and a component of cognition (PC_cognition) based on the Neuropsychological test scores.

| **PC_DSOA** |  | **PC_Cognition** |  |
| --- | --- | --- | --- |
| Variables | Loading | Variables | Loading |
| IPIP-NEO (neuroticism) | 0.733 | MoCA | 0.748 |
| PSS | 0.821 | MMSE | 0.706 |
| HADs anxiety | 0.856 | WLM | 0.702 |
| HADs depression | 0.699 | WLR | 0.761 |
| STAI | 0.852 |  |  |
| TGDS | 0.670 |  |  |
|  | KMO=0.874 |  | KMO=0.613 |
|  | X^2^=314.218 df=15, p<.001, VE=60.1% |  | Χ ^2^ =109.867 (df=6), p <.001 VE=53.2% |

KMO: The Kaiser-Meyer-Olkin Test; VE: Variance explained; Χ^2^: Bartlett’s test of sphericity. ESF, table 1 shows the explanation of neurocognitive test scores and symptoms of anxiety, depression, stress, and personality traits (Tran-Chi et al.,2023).

Abbreviations:

IPIP-NEO: The International Personality Item Pool-NEO (IPIP-NEO), PSS: the Perceived Stress Scale, HADs anxiety: Hospital Anxiety and Depression scale (anxiety), HADs depression: Hospital Anxiety and Depression scale (depression): STAI: the State-Trait Anxiety Inventory (STAI), TGDS: Thai Geriatric Depression Scale, MoCA: the Montreal Cognitive Assessment, MMSE: the Thai-Mini mental state examination, WLM: Word list memory, WLR: word list recognition.

**ESF, Table 2**. Results of binary logistic regression analysis with Neuropsychological test between aMCI and healthy controls (HC)

| **Regression Number** | **Explanatory Variables** | **B** | **SE** | **Wald** | **P** | **OR** | **95% CI** |
| --- | --- | --- | --- | --- | --- | --- | --- |
|  | **Delayed Match to Sample (DMS)** |  |  |  |  |  |  |
| 1 | DMS_MDL | 0.000 | 0.000 | 0.366 | 0.545 | 1.000 | 1.000-1.000 |
| 2 | DMS_PC | -0.038 | 0.021 | 3.062 | 0.080 | 0.963 | 0.923-1.005 |
| 3 | DMS_PEGE | 1.109 | 1.137 | 0.952 | 0.329 | 3.032 | 0.327-28.140 |
| 4 | DMS_PC0 | -0.005 | 0.012 | 0.202 | 0.653 | 0.995 | 0.972-1.018 |
| 5 | DMS_PC4 | -0.018 | 0.010 | 2.926 | 0.087 | 0.982 | 0.962-1.003 |
| 6 | DMS_PC12 | -0.003 | 0.009 | 0.104 | 0.747 | 0.997 | 0.979-1.015 |
| 7 | DMS_PCAD | -0.026 | 0.018 | 2.080 | 0.149 | 0.975 | 0.941-1.009 |
| 8 | DMS_PCS | -0.032 | 0.020 | 2.532 | 0.112 | 0.969 | 0.931-1.007 |
|  | **One Touch Stockings of Cambridge (OTS)** |  |  |  |  |  |  |
| 8 | OTS_MLFC | 0.000 | 0.000 | 0.171 | 0.679 | 1.000 | 1.000-1.000 |
| 9 | OTS_PSFC | **-0.322** | **0.110** | **8.543** | **0.003** | **0.725** | **0.584-0.900** |
|  | **Paired Associates Learning (PAL)** |  |  |  |  |  |  |
| 11 | PAL_TEA | 0.014 | 0.014 | 1.021 | 0.312 | 1.014 | 0.987-1.043 |
| 12 | PAL_FAMS | -0.088 | 0.055 | 2.531 | 0.112 | 0.916 | 0.821-1.021 |
|  | **Pattern Recognition Memory (PRM)** |  |  |  |  |  |  |
| 13 | PRM_PCI | -0.019 | 0.015 | 1.476 | 0.224 | 0.982 | 0.952-1.012 |
| 14 | PRM_PCD | **-0.033** | **0.014** | **5.269** | **0.022** | **0.967** | **0.940-0.995** |
|  | **Reaction Time Task (RTI)** |  |  |  |  |  |  |
| 15 | RTI_FMDMT | 0.001 | 0.003 | 0.282 | 0.595 | 1.001 | 0.996-1.006 |
| 16 | RTI_FMDRT | 0.005 | 0.004 | 1.282 | 0.275 | 1.005 | 0.997-1.013 |
| 17 | RTI_SMDMT | 0.000 | 0.003 | 0.026 | 0.872 | 1.000 | 0.995-1.005 |
| 18 | RTI_SMDRT | 0.004 | 0.004 | 0.874 | 0.350 | 1.004 | 0.996-1.013 |
|  | **Rapid Visual Information Processing (RVP)** |  |  |  |  |  |  |
| 19 | RVP_PFA | 0.004 | 0.002 | 2.696 | 0.101 | 1.004 | 0.999-1.008 |
| 20 | RVP_A´ | **-12.273** | **4.403** | **7.768** | **0.005** | **0.000** | **0.000-0.026** |
| 21 | RVP_MDL | -0.008 | 0.006 | 1.678 | 0.194 | 0.992 | 0.980-1.004 |
|  | **Spatial Working Memory (SWM)** |  |  |  |  |  |  |
| 22 | SWM_BE468 | 0.017 | 0.030 | 0.333 | 0.564 | 1.017 | 0.960-1.079 |
| 23 | SWM_S | 0.057 | 0.148 | 0.147 | 0.701 | 1.058 | 0.792-1.414 |
| 24 | SWM_BE4 | 0.043 | 0.147 | 0.087 | 0.769 | 1.044 | 0.782-1.394 |
| 25 | SWM_BE6 | 0.073 | 0.060 | 1.491 | 0.222 | 1.076 | 0.957-1.210 |
| 26 | SWM_BE8 | -0.021 | 0.047 | 0.205 | 0.651 | 0.979 | 0.892-1.074 |
|  | **Motor Screening Task (MOT)** |  |  |  |  |  |  |
| 27 | MOT_ML | 0.002 | 0.001 | 5.205 | 0.023 | 1.002 | 1.000-1.004 |
| 28 | MOT_SDL | 0.004 | 0.002 | 5.834 | 0.016 | 1.004 | 1.001-1.007 |

**Abbreviations:**

DMS MDL: Delayed Match to Sample median latency

DMS PC: Delayed Match to Sample percent correct

DMS PEGE: Delayed Match to Sample probability of error given error

DMSPC0: Delayed Match to Sample percent correct 0 second delay

DMSPC4: Delayed Match to Sample percent correct 4 second delay

DMSPC12: Delayed Match to Sample percent correct 12 second delay

DMS PCAD: Delayed Match to Sample percent corrects all delay

DMS PCS: Delayed Match to Sample percent correct (Simultaneous)

OTS MLFC: One Touch Stockings of Cambridge median latency to first choice

OTS PSFC: One Touch Stockings of Cambridge probability solved on first choice

PAL TEA: Paired Associates Learning total errors (adjusted)

PAL FAMS: Paired Associates Learning first attempt memory score

PRM PCI: Pattern Recognition Memory percent correct immediate

PRM PCD: Pattern Recognition Memory percent correct delayed

RTI FMDMT: Reaction Time Task median five-choice time

RTI FMDRT: Reaction Time Task median five-choice reaction time

RIT SMDMT: Reaction Time Task simple Median Movement Time

RVP PFA: Rapid Visual Information Processing probability of false alarm

RVP A´: Rapid Visual Information Processing processing A prime

RVP MDL: Rapid Visual Information Processing median response latency

SWM BE468: Spatial Working Memory between errors (4-8 boxes)

SWMS: Spatial Working Memory strategy score (6 or more)

SWMBE8: Spatial Working Memory between errors 8 boxes

MOT ML: Motor Screening Task mean latency

MOT SDL: Motor Screening Task standard deviation of the mean latency

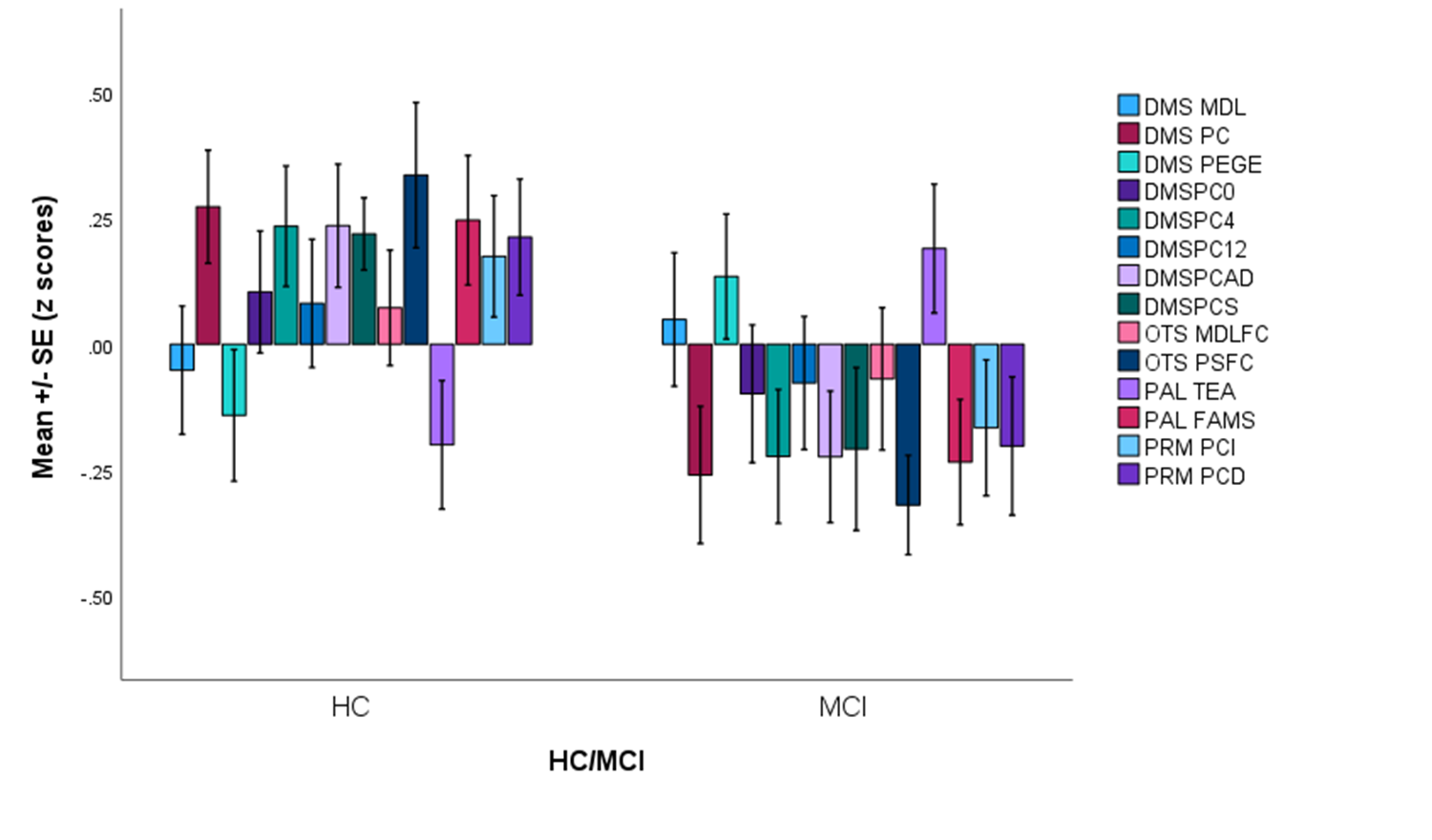

**ESF, Figure 1**. Clustered bar graph showing the z scores of the subdomains of 14 key Cambridge Neuropsychological Test Automated Battery (CANTAB) test scores in amnestic mild cognitive impairment (MCI) versus healthy controls (HC). See ESF, Table 2 for abbreviations.

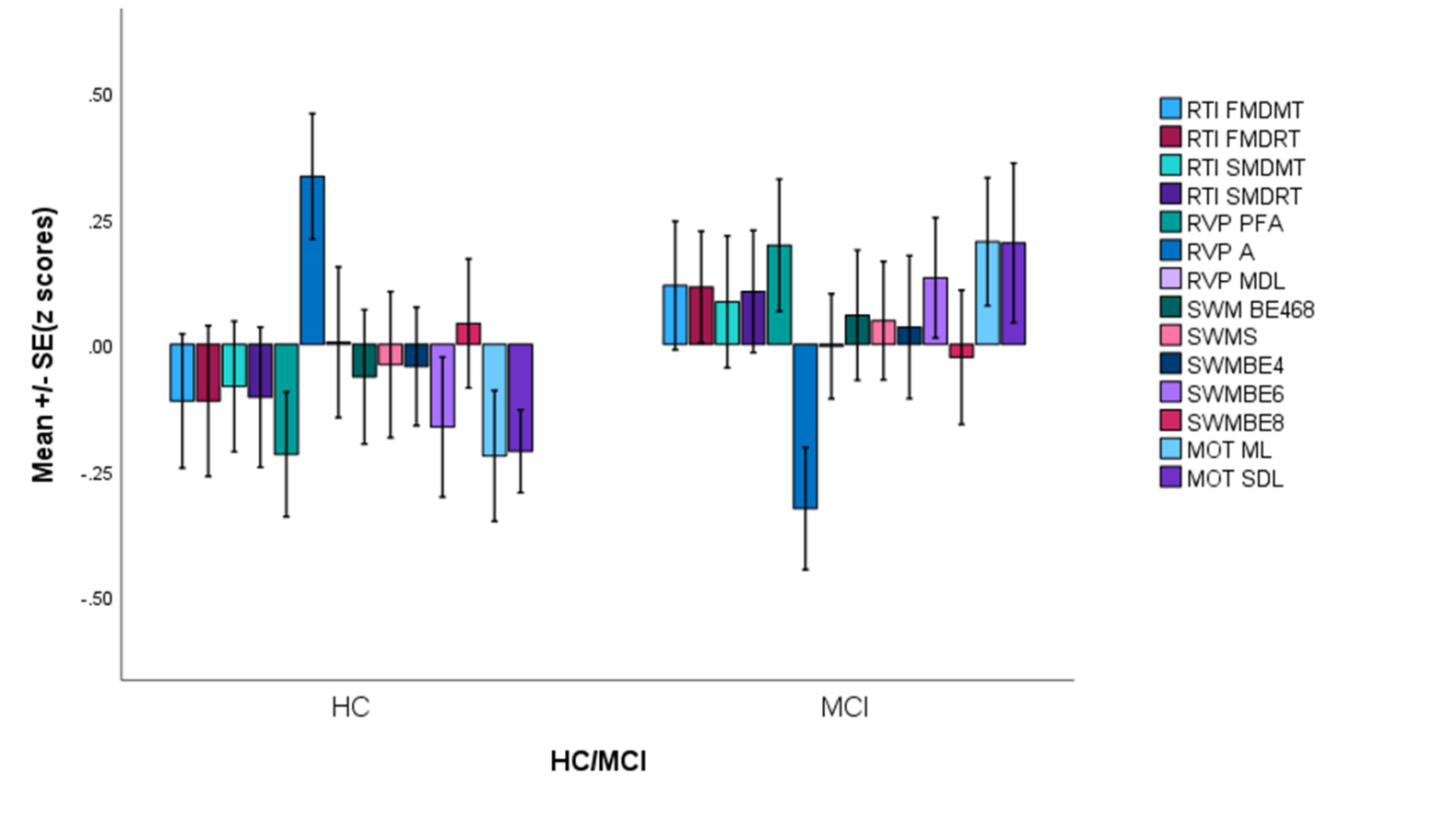

**ESF, Figure 2**. Clustered bar graph showing the z scores of the subdomains of 14 key Cambridge Neuropsychological Test Automated Battery (CANTAB) test scores in amnestic mild cognitive impairment (MCI) versus healthy controls (HC). See ESF, Table 2 for abbreviations.

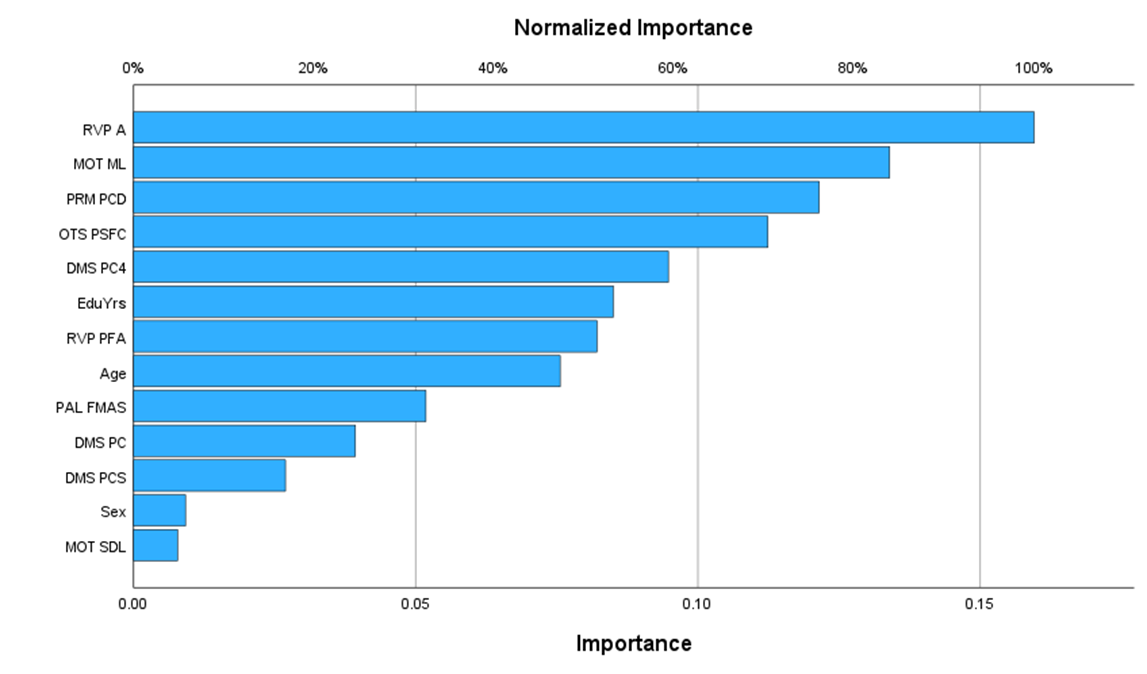

**ESF, Figure 3**. Results of neural network analysis with amnestic mild cognitive impairment and normal controls as output variables, and selected key Cambridge Neuropsychological Test Automated Battery (CANTAB) test scores as input variables. See ESF, Table 2 for abbreviations.
